# Social genomics, cognition, and well-being during the COVID-19 pandemic

**DOI:** 10.1101/2023.05.31.23290618

**Authors:** James R. Bateman, Sudarshan Krishnamurthy, Ellen E. Quillen, Christian E. Waugh, Kiarri N. Kershaw, Samuel N. Lockhart, Timothy M. Hughes, Teresa E. Seeman, Steve W. Cole, Suzanne Craft

## Abstract

**INTRODUCTION:** Adverse psychosocial exposure is associated with increased proinflammatory gene expression and reduced type-1 interferon gene expression, a profile known as the conserved transcriptional response to adversity (CTRA). Little is known about CTRA activity in the context of cognitive impairment, although chronic inflammatory activation has been posited as one mechanism contributing to late-life cognitive decline.

**METHODS:** We studied 171 community-dwelling older adults from the Wake Forest Alzheimer’s Disease Research Center who answered questions via a telephone questionnaire battery about their perceived stress, loneliness, well-being, and impact of COVID-19 on their life, and who provided a self-collected dried blood spot sample. Of those, 148 had adequate samples for mRNA analysis, and 143 were included in the final analysis, which including participants adjudicated as having normal cognition (NC, *n* = 91) or mild cognitive impairment (MCI, *n* = 52) were included in the analysis. Mixed effect linear models were used to quantify associations between psychosocial variables and CTRA gene expression.

**RESULTS:** In both NC and MCI groups, eudaimonic well-being (typically associated with a sense of purpose) was inversely associated with CTRA gene expression whereas hedonic well-being (typically associated with pleasure seeking) was positively associated. In participants with NC, coping through social support was associated with lower CTRA gene expression, whereas coping by distraction and reframing was associated with higher CTRA gene expression. CTRA gene expression was not related to coping strategies for participants with MCI, or to either loneliness or perceived stress in either group.

**DISCUSSION:** Eudaimonic and hedonic well-being remain important correlates of molecular markers of stress, even in people with MCI. However, prodromal cognitive decline appears to moderate the significance of coping strategies as a correlate of CTRA gene expression. These results suggest that MCI can selectively alter biobehavioral interactions in ways that could potentially affect the rate of future cognitive decline and may serve as targets for future intervention efforts.

## Introduction

Alzheimer’s disease (AD) is a progressive neurodegenerative condition that is a common cause of both mild cognitive impairment (MCI) and dementia.^1,2^ Psychosocial risk and resiliency factors can modulate the rate of subsequent cognitive decline in the context of developing neuropathologic changes in the brain, and many of these factors may be modifiable.^3^ Key to developing interventions to harness such resilience effects is identifying the specific psychosocial processes that impact the biology of cognitive decline.

Research in those with normal cognition (NC) has identified some psychobiological pathways through which psychosocial factors may impact biological processes relevant to cognitive function. One such molecular pathway is the conserved transcriptional response to adversity (CTRA).^4^ The CTRA is a pattern of leukocyte gene expression that has been observed across species (i.e., conserved) in response to a host of adverse social conditions. The CTRA transcriptional pattern involves an increase in expression of proinflammatory genes (e.g., *IL1B, IL6, TNF*) and a decrease in the expression of type I interferon response genes (e.g., *IFI-, OAS-* and *MX-*family genes) in response to fight-or-flight stress signaling from the sympathetic nervous system (SNS). A proposed evolutionary explanation for such a response is the necessity to pivot to an anti-microbial, and away from the anti-viral, state of the immune system during periods of acute threat.^5^ Under ancestral conditions, activation of the SNS would have predicted an increased risk of wound-associated bacterial infections and thus support wound healing. In modern conditions, chronic low-level threat produces chronic activation of pro-inflammatory genes, which can contribute to the pathogenesis of a host of common chronic conditions such as neurodegenerative disorders such as Alzheimer’s disease and related dementias,^6^ cardiovascular disease, and neoplastic disorders^7,8^.This chronic, low-grade inflammation also increases with age (“inflammaging”) and the CTRA is one mechanism through which conditions of chronic psychosocial adversity can alter underlying biology and potentially accelerate inflammaging-related disease processes.^9^ However, to date, few studies have evaluated CTRA risk or resilience processes in the context of cognitive aging.

The CTRA was first identified in the context of loneliness,^10^ and subsequent work linked these effects to reduced levels of eudaimonic well-being, or a sense of purpose and meaning in life.^11^ Eudaimonic well-being is distinguishable from hedonic well-being, which is the summation of positive affective experiences in a person’s life.^12^ In the context of cognitive aging, Boyle and colleagues in the Rush Memory and Aging Project (MAP) have previously linked a sense of purpose in life to variations in cognitive aging:^13^ participants with a greater sense of purpose in life were found to have a lower risk of both AD and MCI over a 7-year follow-up period. The biological mechanism through which a sense of purpose relates to cognitive decline is not yet known. However, given the potential role of inflammatory biology in cognitive aging, the CTRA-associated inflammatory biology may represent one mechanism through which psychosocial factors (i.e., a sense of purpose) could affect either risk or resiliency to cognitive decline.

In the present study, we conducted genome-wide transcriptional profiling of dried blood spots collected during a period of significant psychosocial stress - the first 2 years of the COVID-19 pandemic – and examined the links between CTRA gene expression and several psychosocial risk and resilience factors, including loneliness, perceived stress, and both hedonic and eudaimonic well-being. Analyses focused on understanding how the psychosocial correlates of CTRA gene expression may be similar to or different between those with normal cognition and those with mild cognitive impairment.

## Methods

### Participants

All participants were previously enrolled in the Alzheimer’s Disease Clinical Core (Clinical Core) cohort of the Wake Forest Alzheimer’s Disease Research Center (WF ADRC) and underwent standardized evaluations in accordance with the National Alzheimer’s Coordinating Center (NACC) protocol for data collection which meets Uniformed Data Set (UDS) requirements. Specific inclusion and exclusion criteria for the Clinical Core are described elsewhere.^14^ Cognitive adjudication at yearly Clinical Core study visits using clinical and cognitive assessment and brain MRI provides a cognitive diagnosis of NC, MCI, or dementia. For the purposes of this study, only participants with NC or MCI were eligible for study inclusion. The determination of mild cognitive impairment was made using clinical criteria according to NACC guidelines as described by Petersen and Morris.^15^ To address early pandemic concerns of in-person study visits, both questionnaire responses and dried blood spot collection were designed to be collected remotely. All study procedures were approved by the Wake Forest Baptist Health Institutional Review Board. Written informed consent was obtained for all participants and/or their legally authorized representative. Questionnaires were administered via telephone between February 15 and July 21, 2021. Dried blood spot collection occurred a median of 8 days (range: -40 to 136 days) following the questionnaire completion.

### Perceived Stress

The 10-item Perceived Stress Scale (PSS) is a questionnaire that is an index of an individual’s perception of stress over the past month.^16^ Stress was operationalized as finding one’s life unpredictable and uncontrollable, and feeling overloaded. The questions were answered on a Likert scale that ranged from “never” (0) to “very often” (4) after reversing the four positively stated questions. Individual items are summed to produce a total score and showed good internal reliability (α = 0.85). Higher scores reflect higher levels of perceived stress.

### Loneliness

Loneliness was assessed using the UCLA Loneliness Scale, Version 3.^17^ Participants rate statements to describe how often they feel the way described, ranging from “never” (0) to “often” (4). There were twenty statements, and nine were reverse coded according to standard instructions. A total score was computed with higher scores indicating greater feelings of loneliness and showed good internal reliability (α = 0.85).

### Coping

The Brief Coping Orientation to Problems Experienced (brief-COPE) was developed by Carver^18^ as an abbreviated version of the longer COPE, which contains 28 items that measure 14 factors of coping along a Likert scale ranging from “I have not been doing this at all” (0) to “I have been doing this a lot” (3). We added 6 questions related to positive distraction.^19^ Consistent with prior research,^20^ we performed a parallel factor analysis which identified a 3-factor solution of Support (comprised of emotional support, instrumental support, and active coping items), Distraction and Reframing (comprised of positive distraction, positive reframing, and self-distraction), and Blame and Disengagement (comprised of self-blame and behavioral disengagement). Non-participating factors were denial, substance use, venting, planning, humor, religion, and acceptance.

### Hedonic and Eudaimonic Well-Being

The Mental Health Continuum – Short Form (MHC-SF)^12^ is a 14-item questionnaire derived from a 40-item questionnaire.^21^ The MHC-SF was designed to measure hedonic and psychological well-being as conceptualized by Ryff and social well-being as conceptualized by Keyes.^22,23^ Respondents were asked to answer questions about the degree to which they’ve felt a given way over the past month ranging from “never” (0) to “every day (5). Three questions were summed for hedonic well-being (HWB), five for social well-being (SWB), and six for psychological well-being (PWB). SWB and PWB together make up eudaimonic well-being (EWB). The overall internal reliability of the MHC-SF was good (α = 0.89).

### COVID-19 Specific Experiences

Two questionnaires were given to assess the impact of COVID-19 on participants. We administered by telephone the participant version of the COVID-19 Impact Survey, version 1^24^ from the NACC that was initially released in the Summer of 2020. This questionnaire captured information regarding COVID-19 exposure, medical consequences, impact on psychosocial factors, and perceived cognitive, psychiatric, and behavioral consequences. We supplemented this questionnaire with questions from the Questionnaire for Assessing the Impact of COVID-19 Pandemic and Accompanying Mitigation Efforts on Older Adults (QAICPOA).^25^ The QAICPOA was developed specifically to assess the impact of COVID-19 on older adults and includes questions regarding diagnosis, symptoms, actions taken because of the pandemic, changes in contact and communication. Some questions included in these forms were discrete dates (e.g., dates of COVID diagnosis), others were yes/no responses to questions regarding types of care accessed. For the purposes of this study, three questions were included in the analysis, all from the COVID-19 Impact Survey. Questions 7 (worry about COVID-19 infection/reinfection), 8 (isolated or cut off from family and friends due to COVID-19), and 9 (disruption to everyday life due to COVID-19). Each of these questions were answered on a five point Likert scale ranging from “not at all” (one) to extremely (five).

### Dried Blood Spot Collection

After questionnaires were collected, participants were mailed a remote collection kit for self-collection of dried blood spots. Training materials were adapted for use in our cohort from Allen and colleagues.^26^ Participants were sent all necessary materials and a printed instruction booklet with instructions on specimen collection. Blood spots were placed directly on a standardized filter paper commonly used for neonatal screening (Whatman #903, GE Healthcare, Piscataway, NJ). Pictorial examples of both good and bad dried blood spot collection were provided. Participants were instructed to allow the collection card to sit for four hours to dry, then place the folded collection card, along with a humidity detector and silica gel packs, in a provided gas permeable bag and return to the WF ADRC in a provided return envelope.

### Measurement of Gene Expression

Dried blood spot were stored at -80°C at the WF ADRC and then shipped as a single batch on dry ice to the UCLA Social Genomics Core Laboratory for transcriptome-wide RNA profiling and CTRA gene expression analyses as previously described.^27,28^ Briefly, RNA was extracted (Qiagen RNeasy), converted into cDNA using a high-efficiency mRNA-targeted reverse transcription system (Lexogen QuantSeq 3’ FWD), and sequenced on an Illumina NovaSeq instrument in the UCLA Neuroscience Genomics Core Laboratory, all following the manufacturers’ standard protocols for this workflow. Sequencing targeted >10 million single stranded 100-nt reads per sample (achieved median = 17.3 million), each of which was mapped to the GRCh38 reference human transcriptome using the STAR aligner (median 83% mapping rate), and quantified as gene transcripts per million total mapped reads with expression values floored at 1 transcript-per-million to suppress spurious low-range variability, log2-transformed to stabilize level-dependent variance within gene, and z-score transformed to stabilize variance across genes. Among 171 assayed samples, routine post-assay data quality screening identified 7 samples with insufficient RNA sequencing reads (< 5 million), 8 additional samples with poor read mapping rates (< 70%), and 6 additional samples with poor signal-to-noise ratios (average profile correlation with other samples: r < .50), leaving a total of 150 valid RNA profiles available for analyses of CTRA. This 88% valid data yield is consistent with previous research involving genome-wide transcriptional profiling of dried blood spot samples.^27,28^

### Statistical Analysis

As in previous dried blood spot CTRA studies, we used linear mixed effect models to analyze average expression of a pre-specified set of CTRA indicator gene transcripts as a function of psychosocial risk and resilience factors while controlling for covariates. Analyses focused on a pre-specified set of 53 CTRA indicator genes used in previous research,^4,29^ of which 43 were reliably detectable in this study, including 16 pro-inflammatory gene transcripts (*CXCL8, FOS, FOSB, FOSL2, IL1B, JUN, JUNB, JUND, NFKB1, NFKB2, PTGS1, PTGS2, REL, RELA, RELB, TNF*) and 27 Type I interferon-related gene transcripts (*GBP1, IFI16, IFI27, IFI27L2, IFI35, IFI44, IFI44L, IFI6, IFIH1, IFIT1-IFIT3, IFIT1B, IFIT5, IFITM1-IFITM3, IRF2, IRF7, IRF8, JCHAIN, MX1-MX2, OAS1-OAS3, OASL*), and 10 of which were removed due to minimal expression levels or variation (SD < .5 log2 expression units; *FOSL1, IFI27L1, IFI30, IFITM4P, IFITM5, IFNB1, IGLL1, IGLL3P, ILA1, IL6*). Gene-specific z-score signs were reversed for the antiviral gene set to reflect its inverse contribution to the CTRA profile.^4^ Mixed models were estimated by maximum likelihood (SAS PROC MIXED) and specified fixed effects of indicator gene (repeated measure), cognitive status (normal vs mild cognitive impairment), psychosocial risk/resilience factors, a cognitive status x psychosocial factor interaction term (testing for differences in CTRA association as a function of cognitive status), and covariates (age, sex, race, BMI, history of regular smoking, and history of regular heavy alcohol consumption); a random effect of study participant; and a fully saturated (unstructured) variance-covariance matrix to account for residual heteroscedasticity and correlation across participants. In the event of a significant cognitive status x psychosocial factor interaction, additional follow-up “simple slopes” analyses quantified the association of psychosocial factors with CTRA gene expression nested within cognitive status group.

## Results

### Demographics and Cognitive Status

A total of 171 participants provided dried blood spot samples (106 NC, 58 MCI, 1 Dementia, 6 Other/NA) with 148 of those samples (87%) yielding valid RNA data. Participants without diagnoses of NC or MCI were excluded from further analysis (4), and 1 participant completed dried blood spot without questionnaires and was excluded from further analysis, yielding a final analytic sample of 143 participants: 91 with normal cognition and 52 with MCI. The mean age of our group was 72.9 +/- 8.04 years, 16% were Black, and 69% were female, and 18% were treated with a beta-blocker. Participant demographic characteristics are summarized in Table 1. Less than 4% of data were missing for all variables analyzed.

**Table 1.**
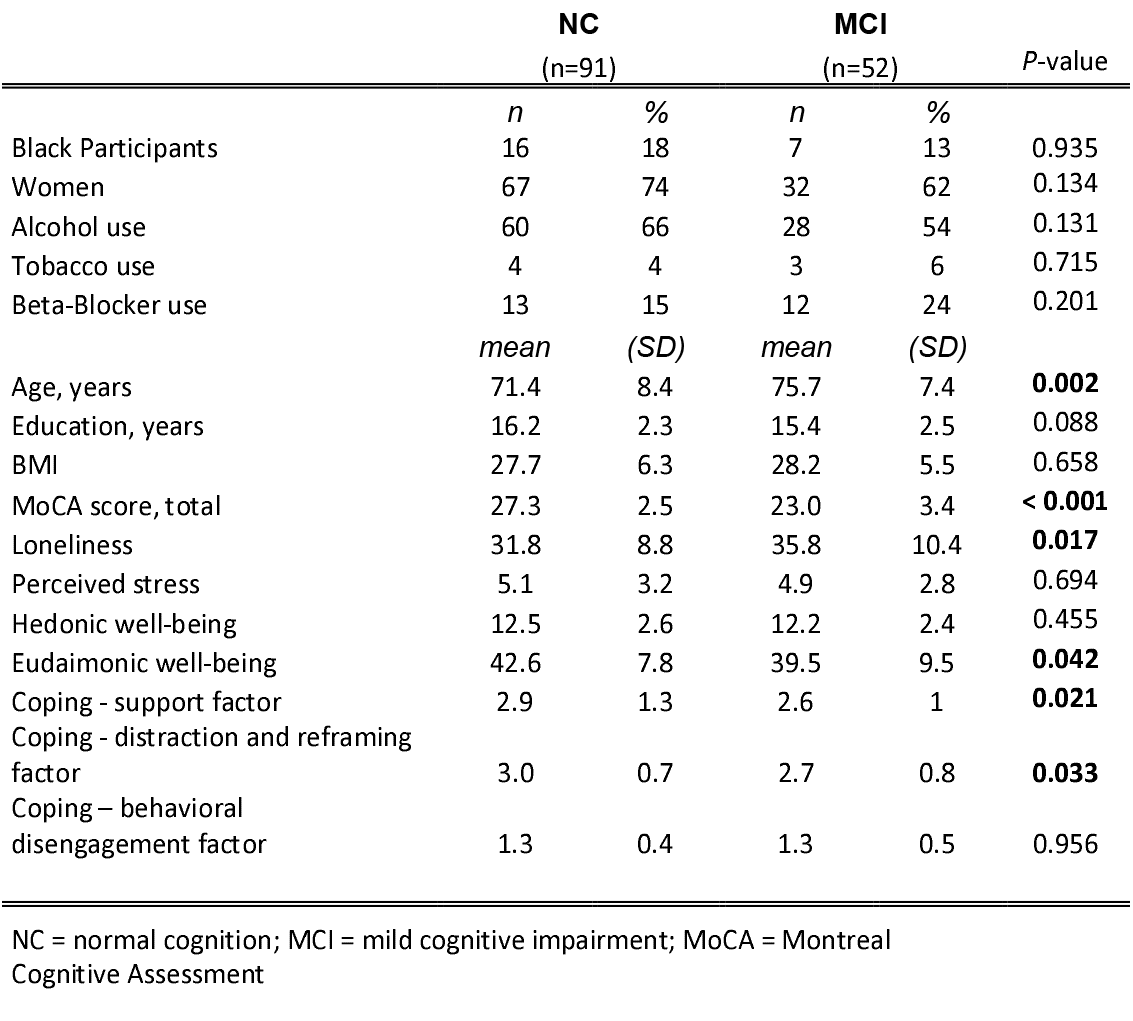
Demographics.

### Cognitive Impairment and the Psychosocial Correlates of CTRA

To determine how cognitive impairment might affect the relationship between psychosocial factors and CTRA gene expression, we compared the relation of CTRA gene expression to psychosocial risk factors (stress, loneliness), two distinct domains of well-being (hedonic and eudaimonic well-being), and three distinct domains of coping (blame and disengagement, distraction and reframing, and social support) for NC and MCI groups while controlling for covariates.

Consistent with previous reports,^11,30,31^ CTRA gene expression was significantly associated with the 2-dimensional representation of well-being (distinct hedonic and eudaimonic dimensions; F(2, 129) = 4.93, *p* = 0.009; Table 2, Model 1), with a significant inverse association with eudaimonic well-being (−0.045 log_2_ RNA per well-being SD ± 0.015 SE, *p* = 0.003). There was no significant association with hedonic well-being (+0.019 ± 0.015, *p* = 0.225). Neither stress nor loneliness showed any significant association with CTRA gene expression in this sample (F(2, 130) = 0.37, *p* = 0.693; individual *p*’s > 0.50; Table 2, Model 2).

**Table 2.**
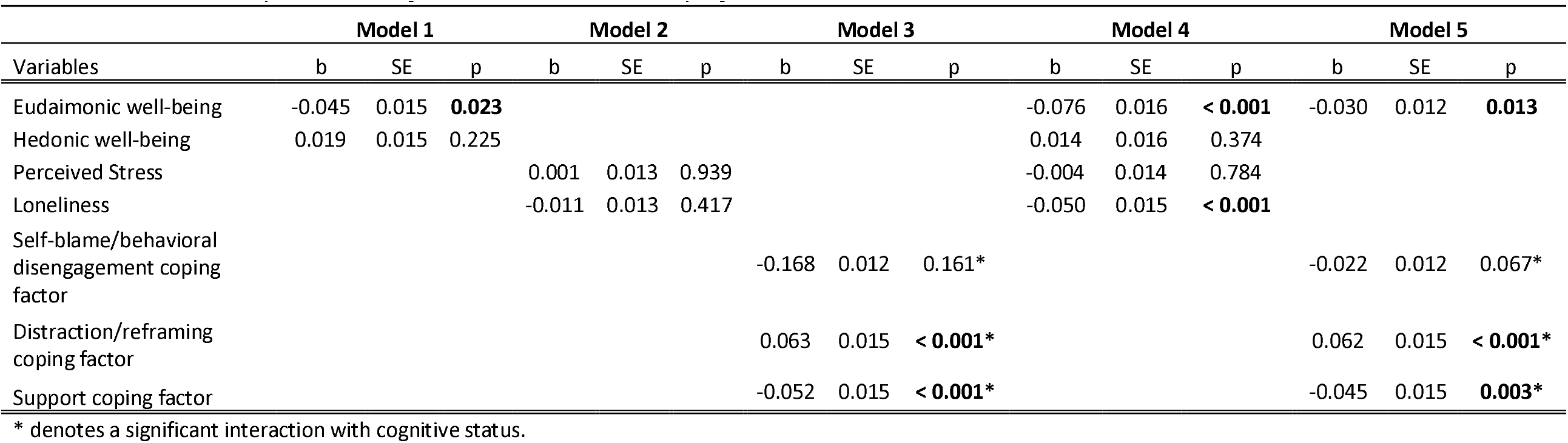
CTRA relationship to well-being, stress, loneliness, and coping factors.

CTRA gene expression also varied significantly as a function of the three major dimensions of coping in this sample (F(3, 126) = 7.22, *p* < 0.001; Table 2 Model 3, Figure 1). However, we detected significant interactions between cognitive status and the brief-COPE as it relates to CTRA gene expression (F(3, 123) = 9.07, *p* < 0.001). Among those with normal cognitive function, coping through social support was associated with lower CTRA gene expression (−0.075 ± 0.017, *p* < 0.001) whereas coping by distraction / reframing was associated with higher CTRA gene expression (+0.086 ± 0.018, *p* < 0.001). Among those with MCI, coping by blame or disengagement was associated with a lower CTRA gene expression (−0.077 ± 0.018, *p* < 0.001).

**Figure 1.**
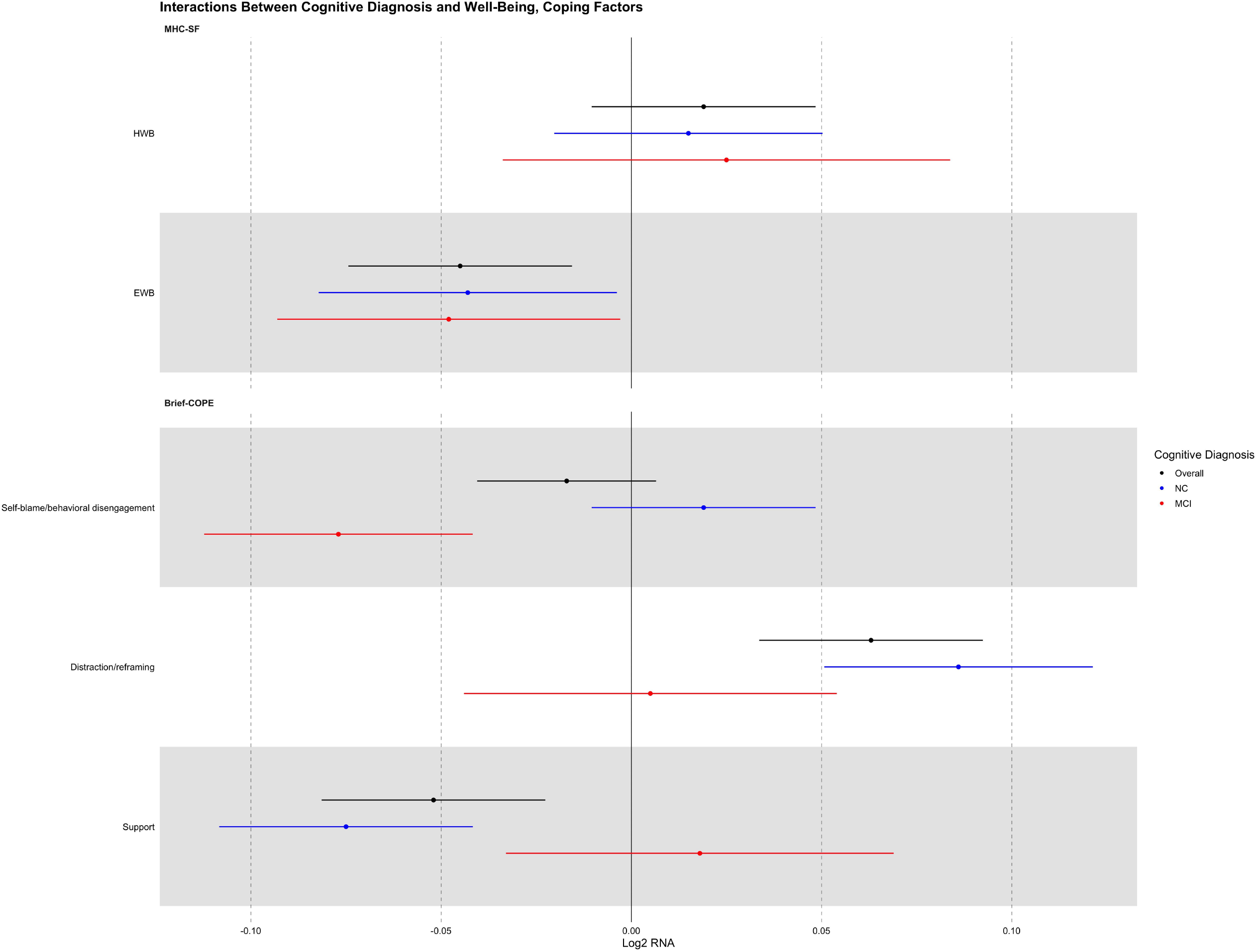
Interactions between cognitive diagnosis and well-being, coping factors. Forest plot demonstrating the strength of association (b ± SE) between the indicated predictor variable and the 53-gene CTRA contrast score for two dimensions of well-being and three coping factors.

CTRA gene expression was significantly associated in a model containing both psychosocial risk factors (perceived stress, loneliness) and dimensions of well-being (eudaimonic, hedonic); F(2, 126) = 11.72, *p* < 0.001; Table 2, Model 4), with a significant inverse relationship between CTRA gene expression and eudaimonic well-being (−0.076 ± 0.016, *p* < 0.001) and loneliness (−0.050 ± 0.015, *p* < 0.001) with no effect modification by cognitive diagnosis. There are significant correlations between eudaimonic well-being and hedonic well-being (R = 0.60), loneliness (R = -0.59), and perceived stress (R = -0.42).

A model including both eudaimonic well-being and the three coping factors was significantly associated with CTRA gene expression; F(3, 123) = 7.01, *p* < 0.001 (Table 2, Model 5). In this model, there was a significant inverse correlation between CTRA gene expression and eudaimonic well-being (−0.03 ± 0.012, *p* = 0.013) with no effect modification of cognitive diagnosis. However, we again detected significant interactions between cognitive status and the brief-COPE in relation to CTRA gene expression (F(3, 119) = 10.71, *p* < 0.001). Among those with normal cognitive function, coping through social support was associated with a lower CTRA gene expression (−0.076 ± 0.017, *p* < 0.001) whereas coping by distraction / reframing was associated with a higher CTRA gene expression (+0.091 ± 0.017, *p* < 0.001). Among those with MCI, coping with blame or disengagement was associated with a lower CTRA gene expression (−0.080 ± 0.018, *p* < 0.001). There was no significant correlation between eudaimonic well-being and these coping factors.

Several COVID-19-related factors were associated with significant differences in CTRA gene expression, none of which differed by cognitive diagnosis (F(3, 124) = 3.20, p = 0.026). A past diagnosis of COVID-19, either confirmed or suspected, was associated with a lower CTRA gene expression (−0.144 ± 0.058, *p* = 0.014). Of note, only 6 out of 143 (4%) of our participants reported a past COVID diagnosis, and the timing of past infection was not documented. Participants who reported feeling isolated due to COVID-19 had a lower CTRA gene expression (−0.027 ± 0.011, *p* = 0.019), and those who reported higher distress had a higher CTRA gene expression (+0.030 ± 0.018, *p* = 0.019). Degree of worry about COVID-19 was not significantly associated with CTRA gene expression (+0.024 ± 0.015, *p* = 0.105).

## Discussion – 1522 need to remove 22 words…

Our analysis of genome regulation in the context of the COVID-19 pandemic during the period of general social distancing documented distinctive transcriptional correlates of well-being and dimensions of coping. In both MCI and NC, these data are consistent with previous research in identifying an inverse association of CTRA gene expression with eudaimonic well-being. For the NC group, CTRA gene expression was also inversely associated with coping through social support, but directly (unfavorably) associated with coping by distraction and reframing. By contrast, CTRA gene expression was not associated with either of those coping dimensions for individuals with MCI. The patterns of similar and distinct associations for MCI vs NC suggests that broad experiences of psychological and social well-being remain centrally relevant to biobehavioral function in the context of MCI, whereas more specific dimensions of self-management and coping may become less relevant to individual biobehavioral function in the context of MCI as individuals come to depend more on others to help support activities of daily life and cope with challenge, and thus less predominately dependent on their own cognitive processes and coping responses.

High levels of loneliness have been shown to be associated with an upregulation of CTRA gene expression.^32^ However, loneliness did not predict CTRA profile in our cohort. It is possible that the relatively low loneliness scores among our participants was below a threshold at which an effect would be seen. One previous study found that eudaimonic well-being had a stronger relationship with CTRA gene expression than did loneliness when both variables are considered simultaneously, suggesting that the two variables’ effects may stem from their common involvement in social well-being.^33^ In a model containing both eudaimonic well-being an loneliness, we found that eudaimonic retained a significant inverse association with CTRA gene expression but a counterintuitive inverse relationship between loneliness and CTRA gene expression appeared after the shared variance between these two variables of interest was accounted for, a finding that will need to be explored in future work. Previous studies of the association of eudaimonic and hedonic well-being with CTRA profiles have demonstrated similar findings to that of our cohort. In a study of 84 healthy adults aged 35 – 64, eudaimonic well-being was associated with downregulated CTRA gene expression, while hedonic well-being was associated with CTRA upregulation.^30^

Wyman and colleagues^34^ recently reported on the association between psychological well-being as measured in the NIH Toolbox on Emotion and found that, in a race-stratified analysis, Black and American Indian/Alaskan Native participants reported lower life satisfaction than White participants, but similar scores on positive affect, meaning in life, and purpose in life. Measures of executive functions, but not episodic memory, were higher in those with higher life satisfaction scores. Psychological well-being is a multi-dimensional construct, and includes evaluative well-being related to evaluations made about life, hedonic well-being or pleasures and satisfaction from life, and eudaimonic well-being or a sense of greater purpose in life.^35^ Subjective clinical complaints associated with MCI, such as memory concerns, are predictive of reduced psychological well-being in individuals.^36^ Additionally, neuropsychiatric symptoms in MCI are associated with increased risk of incident dementia, independent of prior functional or cognitive status.^37^ It is possible that subjective clinical complaints and neuropsychiatric symptoms associated with MCI lead to the observed reduction in eudaimonic well-being among MCI participants in this study. Interventions targeting subjective self-reported health and emotional factors related to well-being have the potential to improve eudaimonic well-being and reduce the associated upregulation in CTRA profile.

The default mode network, an intrinsic connectivity network that is central to AD pathogenesis,^38^ has been implicated in the neural correlates of loneliness and a sense of meaning and purpose in life (a component of eudaimonic well-being).^39^ Internetwork connectivity was more dense and less modular between the default mode network, and the frontoparietal, attention, and perceptual networks in lonely individuals. Conversely, a greater sense of meaning in life was associated with an increase in modularity between the default mode and limbic networks. This work suggests that the default mode network is a central hub that is involved in shifting between states through differences in modularity and integration with the frontoparietal and limbic networks. These data suggest that the benefits of eudaimonic well-being translate to older populations with normal cognition as well as those with MCI. Interventions shown to improve eudaimonic well-being and reduce CTRA gene expression, such as mindfulness meditation practices,^31^ might be investigated in future studies with NC and MCI individuals to gauge improvements in eudaimonic well-being and corresponding CTRA profiles.

Strategies used for coping with psychosocial stress in MCI have been assessed in prior work, though this is the first study to evaluate its molecular correlates in gene expression. Coin and colleagues^40^ assessed coping strategies in people living with MCI and dementia. This study utilized the COPE, but used *a priori* categories of coping, including five scales of problem-focused coping, five scales of emotion-focused coping, and three scales of coping strategies sometimes considered less healthy (venting of emotions, behavioral disengagement, and mental disengagement). They found that individuals with greater cognitive impairment had poorer coping strategies. This association remained present even after adjusting for pre-pandemic depression, suggesting that less efficient coping strategies may have exposed those with a greater degree of cognitive impairment to more psychosocial stress related to pandemic-related social distancing. Murukesu and colleagues^41^ assessed coping strategies using the brief-COPE in older adults with cognitive frailty during the period of restricted movement, travel, and assembly in Malaysia which looked at well-being and coping between two groups in a randomized control trial of a multi-domain intervention to address cognitively frailty. They found that older adults with cognitive frailty used religion, acceptance, and positive reframing (i.e., active coping) and self-blame, denial, and substance use (i.e., avoidant coping) were the least common. Our study adds to this literature in defining the molecular correlates of coping in the context of immune cell gene expression.

We also did not utilize *a priori* assumptions of coping strategies and rather relied on a data-driven approach to evaluate coping in our cohort. In our sample, we found that only those with normal cognition demonstrated a reduction in CTRA gene expression with the use of social support. At the same time, among those with normal cognition, coping based on distraction and reframing was associated with elevated CTRA gene expression. One possibility that is beyond the scope of our data to answer is that, as individuals develop prodromal cognitive decline (i.e., MCI), self-appraised coping strategies may become less clearly associated with actual coping strategies. A similar pattern is seen in the self-appraisal of cognitive impairment, where those with MCI demonstrate a progressive underappreciation of their own cognitive deficits.^42^ Given that possibility, it becomes all the more remarkable that eudaimonic well-being remains an important correlate of molecular well-being for adults with MCI as well as for adults with normal cognition, and that consummatory sources of hedonic wellbeing remain a risk even in the context of MCI. Again, this pattern could potentially reflect that persons with MCI may be “outsourcing” their coping to their caregivers/support network, and thus their own psychological reactions bear little relationship to their CTRA biology whereas their engagement with others (social well-being) is the primary psychosocial source of biological resilience.

Several issues limit the interpretation of the present results. In this sample, people with cognitive impairment were both older and lonelier than those without cognitive impairment, and this range-restriction could have contributed to the lack of association observed for loneliness and CTRA gene expression among those with MCI. The present results come from data collected during the COVID-19 pandemic and associated social distancing protocols, and it is unclear whether similar results would be obtained in other settings and social conditions. Our MCI sample was smaller than our NC sample, potentially leading to asymmetric power across subgroups. Because our data come from a single regional context, it is unclear whether our findings would hold true across all individuals with MCI, and future work should focus on larger and more broadly representative samples.

Despite these limitations, our work demonstrates several important findings. This is the first study to demonstrate the similar transcriptional correlates of eudaimonic vs. hedonic well-being in individuals with MCI compared to NC individuals. It is well-established that individuals with greater eudaimonic well-being (i.e., a sense of purpose in life) demonstrate a reduced CTRA gene expression profile,^30,31,33^ and past work has found a significant reduction in the risk of AD and MCI associated with a greater sense of purpose in life.^13^ The findings here suggest one potential mechanism through which this psychological resiliency factor may function, mediating a lower inflammatory burden and protecting against the “inflammaging” that has been proposed to contribute to the AD neuropathological cascade.

## Data Availability

All data are available upon reasonable request to the authors and via www.wakeshare.org

## Acknowledgements

Dr. Bateman is supported by the Alzheimer’s Association (AACSF-21-852529, Dementia Alliance of North Carolina, and the NIH (P30AG072947, P30AG049638). Dr. Bateman has received an honorarium from the North Carolina Neurological Society. Since Dr. Bateman is an employee of the U.S. Government and contributed to this manuscript as part of his official duties, the work is not subject to US copyright. This research is supported by the Department of Veterans Affairs Office of Academic Affiliations Advanced Fellowship Program in Mental Illness Research and Treatment and the Department of Veterans Affairs Mid-Atlantic Mental Illness Research, Education, and Clinical Center (MIRECC).

## Conflicts of Interest

Dr. Bateman reports no conflicts of interest.

Mr. Krishnamurthy reports no conflicts of interest.

Dr. Quillen reports no conflicts of interest.

Dr. Waugh reports no conflicts of interest.

Dr. Kershaw reports no conflicts of interest.

Dr. Hughes reports no conflicts of interest.

Dr. Seeman reports no conflicts of interest.

Dr. Cole reports no conflicts of interest.

Dr. Craft reports no conflicts of interest.

## Notes

### Competing Interest Statement

The authors have declared no competing interest.

### Funding Statement

This study was supported by NIH P30AG072947 and P30AG049638.

### Author Declarations

The IRB of Wake Forest University School of Medicine (IRB00025540) gave ethical approval for this work as part of the Alzheimer's Disease Clinical Core Cohort

### Summary of Updates

Fixed misspelling of coauthor name (Krishnamurthy)

